# Associations between Elevated Circulating Amino Acid Levels and Brain Health: A Mendelian Randomization Study in the UK Biobank

**DOI:** 10.64898/2026.07.13.26357918

**Authors:** Andrew C. Mason, Caroline E. Dale, Reecha Sofat, Nishi Chaturvedi, Victoria Garfield

## Abstract

**Background:** previous research has demonstrated a relationship between chronically elevated postabsorptive amino acid (AA) levels and the pathophysiology of neurological disorders such as dementia. It is unclear if some of these associations are causal or secondary to other pathologies. Amino acid metabolism also has a complex relationship with glycaemia, which itself may be implicated in brain health and the aetiology of dementia.

**Study design:** we implemented a two-sample Mendelian randomisation (MR) in the UK Biobank (maximum *N*=375,078 participants) utilising exposure data from a recent genome-wide association study (GWAS) of circulating metabolites. Both univariate and multivariable MR methods were implemented, complemented by standard sensitivity analyses using the weighted median estimator, Egger regression, and MR-PRESSO. Our exposures were genetic instruments proxying elevated levels of the branched chain AAs (BCAAs; isoleucine, leucine, and valine), aromatic AAs (AAAs; phenylalanine, tyrosine, and histidine), and glycated haemoglobin A1c (HbA1c). Our outcomes were subcortical volume measures (of the accumbens, amygdala, caudate, hippocampus, pallidum, putamen, and thalamus); total brain and white matter hyperintensity (WMH) volumes; and all-cause dementia (ACD), vascular dementia (VaD), and Alzheimer’s dementia (AD).

**Results:** we found evidence of direct associations independent of glycaemia between genetically-proxied elevated circulating tyrosine and multiple subcortical volumes – of the accumbens (β = 11.0 95%CI [4.2, 17.8] mm^3^ / mmol L^-1^), caudate (β= 30 95% CI [2.9, 5.71] mm^3^ / mmol L^-1^), hippocampus (β= 43.1 95%CI [14.1, 72.1] mm^3^ / mmol L^-1^), and thalamus (β= 81.4 95%CI [8.54, 154.3] mm^3^ / mmol L^-1^). For dementia outcomes, elevated valine and histidine were associated with a reduced (OR = 0.45 95% CI [0.21, 0.96] / mmol L^-1^) and increased (OR = 1.47 95% CI [1.03, 2.09] / mmol L^-1^) vascular dementia risk, respectively.

**Conclusions:** elevated circulating tyrosine was associated with increases in several subcortical volumes, and these effects were found to be independent of glycaemia. This warrants further investigation as to the effects and possible benefits of tyrosine modification or supplementation in the diet to protect against brain atrophy and the development of neurological disorders. On the other hand, associations between circulating AAs and vascular dementia may indicate mechanistic effects of chronically elevated AA levels on dementia.

## Introduction

The large neutral amino acids (LNAAs) include the branched-chain (BCAAs; isoleucine, leucine and valine) and aromatic amino acids (AAAs; phenylalanine, histidine, and tryptophan) and are obtained from dietary protein. In the postabsorptive state, chronically elevated fasting plasma LNAA concentrations have been linked to dyslipidemia, insulin resistance, impaired glycaemia, along with the onset and progression of type 2 diabetes mellitus (T2DM) (Coker & Coker 2025; Newgard et al. 2009). Given the established links between metabolic dysregulation and neurological disease, understanding whether these metabolites independently affect brain health is of clinical importance.

Altered AA metabolism can negatively impact brain function. For example, maple syrup urine disease (MSUD; a rare inborn error [IEM] of BCAA metabolism) can cause severe neurological impairment (Hassan & Gupta 2026; Haymond, Ben-Galim & Strobel 1978; Siddik et al. 2022; Snyderman et al. 1984; Xu, Jakher & Ahrens-Nicklas 2020). Because BCAAs and AAAs compete for the same large amino acid transporter (LAT-1) to cross the blood brain barrier (BBB), elevated BCAA levels characteristic of MSUD can disrupt CNS LNAA homeostasis (Brosnan & Brosnan 2006). AAAs are the precursors for neurotransmitters such as dopamine, serotonin, and melatonin. Thus, peripheral BCAA elevation results in the depletion of these neurotransmitters. By contrast, phenylketonuria (PKU; a rare inborn error of phenylalanine metabolism) is also associated with neurological impairment as chronically elevated phenylalanine competitively blocks the uptake of other LNAAs (Jaulent et al. 2020; Velema et al. 2015). Acute changes in amino acid levels, such as in the documented onset of IEMs in previously functioning adults (Tufekcioglu et al. 2016), can quickly lead to similar neurological outcomes. Altered serum and CSF amino acid profiles also feature in neurological disorders (such as Alzheimer’s and Parkinson’s disease [AD, PD]), but to date it is not known if these are causal or secondary to pathology (Aquilani et al. 2023; Hernandez et al. 2025; Siddik et al. 2022; Tynkkynen et al. 2018). Observational studies alone cannot yield answers due to confounding and reverse causation; thus, research has turned to leveraging genetic data to investigate causal associations between metabolites and clinical outcomes.

There is evidence from Mendelian randomisation (MR) studies for a causal association between circulating amino acid levels and impaired glycaemia (Lotta et al. 2016; Yuan & Larsson 2020; Zhou et al. 2026). However, some bidirectional analyses suggest that chronically elevated amino acid levels may rather be secondary to T2DM and insulin resistance (Jia et al. 2025). Impairments of amino acid metabolism and glycaemia have separately been associated with neurological disorders (Mason et al. 2026; Tynkkynen et al. 2018). Thus, here we used MR to investigate associations between elevated amino acid levels and brain health, indexed by total brain and clinically relevant subcortical volumes, as well as dementia status. We implemented multivariable MR methods including a genetic instrument for glycaemia indexed by increasing HbA1c levels to constrain whether associations between amino acid levels and brain health are mediated by or independent of glycaemia.

## Materials and methods

### Study design

Two-sample Mendelian randomisation was implemented. We included standard sensitivity analyses to identify potential unbalanced (directional) horizontal pleiotropy (detailed below in *Two sample MR analyses*).

### Sample

Full details of the UK Biobank cohort are published elsewhere (Sudlow et al. 2015). Briefly, UKB comprises ∼500,000 males and females from the general UK population, aged 40-69 years at baseline (2006-2010). There was a maximum of *N* = 375,078 participants with European ancestry that had both genotype and outcomes of interest (see Table 1) available. The UKB received ethical approval from the Northwest Multicentre Research Ethics Committee and informed consent was obtained from participants.

**Table 1:** Sample characteristics for UKB participants considered in our study (n = 375,078)

| Sample characteristic | Mean (SD) / N (%) / Median (IQR) |
| --- | --- |
| Age (years) | 56.9 (8.0) |
| Sex (Female) | 205730 (54.1%) |
| Socioeconomic_deprivation score | -1.56 (2.93) |
| Smoking | ☐ |
| *Ever* | 93916 (27%) |
| Physical activity (number of days) | 3.6 (2.3) |
| Hippocampal volume (mm<sup>3</sup>) | 3790.0 (449) |
| White matter hyperintensity volume (mm<sup>3</sup>)<br>* | 3063 (4680) |
| Body Mass Index (kg/m<sup>2</sup>) | 27.1 (4.8) |
| Glycated haemoglobin (HbA1c;<br>mmol mol<sup>-1</sup>) | 36.0 (6.5) |
| Type 2 diabetes mellitus | 32,970 (8.8%) |
| All-cause dementia | 8,261 (2.2%) |
| Vascular dementia | 1,789 (0.5%) |
| Alzheimer's dementia | 3,726 (1.0%) |
| Isoleucine (mmol mol<sup>-1</sup>) | 0.05 (0.02) |
| Leucine (mmol mol<sup>-1</sup>) | 0.10 (0.03) |
| Valine (mmol mol<sup>-1</sup>) | 0.21 (0.04) |
| Tyrosine (mmol mol<sup>-1</sup>) | 0.06 (0.01) |
| Histidine (mmol mol<sup>-1</sup>) | 0.07 (0.01) |
| Phenylalanine (mmol mol<sup>-1</sup>) | 0.05 (0.01) |
| *Median (IQR) |  |

### Genotyping and quality control in UKB

At the outset, UKB genotyped 487,409 participants using one of two customised genome-wide arrays that were imputed to a combination of the UK10K, 1000 Genomes Phase 3, and the Haplotype Reference Consortium reference panels, which resulted in 93,095,623 autosomal variants. We applied additional variant-level QC, excluding variants with a low call rate (<5%), a minor allele frequency <5%, those that deviated from Hardy-Weinberg equilibrium (HWE; *p<*5e-7), and any multi-allelic or palindromic variants. At the individual-level, we excluded any participants with excessive or minimal heterozygosity, more than 10 putative third-degree relatives derived from the kinship matrix, no consent to extract DNA, mismatches between self-reported and genetic sex, missing QC information, and non-European ancestry. European ancestry was assessed by comparing self-reported ancestry and genetic ancestry derived from principal component analysis of their genotype (Bycroft et al. 2017). If these agreed, they were included. After performing these steps, the sample size was *N*=375,078 participants.

### Variables

#### Exposures SNPs

##### Elevated circulating amino acid levels

Genome-wide significant (*p*<5×10^-8^) SNPs indexing increased levels of individual circulating BCAAs and AAAs were selected from a recent GWAS (Karjalainen et al. 2024). Chosen AAs were isoleucine, leucine, valine, phenylalanine, tyrosine, and histidine. All circulating amino acid measurements have units mmol L^-1^. A pragmatic approach to instrument selection was adopted (see **Genetic instrument selection**).

##### Circulating HbA1c

SNPs indexing increased HbA1c were selected from the European component of the trans-ancestral MAGIC consortium GWAS (Chen et al. 2021), where the data were limited to only those with genome-wide significance (*p*<5×10^-8^). HbA1c has units mmol mol^-1^. Linkage disequilibrium (LD) thresholding to select the final instrument is described below and was performed using the same method applied to individual amino acids.

#### Genetic instrument selection

For all instruments, linkage disequilibrium clumping was performed. Rather than adopting the standard combination of the correlation coefficient threshold and window size in kilobases (r^2^=0.001, *kb*=10,000, respectively), we adopted a more pragmatic approach. Using the 1000G CEU (Utah residents with Northern and Western European ancestry) cohort as a reference panel, we constructed a grid of input R^2^ and *kb* and performed LD clumping using PLINK 2.0 for every combination. Skew and kurtosis were determined from the distributions of exposure data to assess the need for additional transformations, which were not necessary. For the lead SNPs identified for every combination, we computed the association between these SNPs and the relevant exposure from the UKB data (excluding measurements 3-SDs from the mean) via linear regression. On an exposure-by-exposure basis, the adopted instrument was taken as the list of lead SNPs identified that had both the highest proportion of variance explained (R^2^) and F-statistic.

#### Radiomic outcomes: subcortical, total brain, and white matter hyperintensity volumes

Brain MRI images were acquired using a Siemens Skyra 3-T scanner with a standard Siemens 32-channel head coil. We utilized imaging-derived phenotypes derived from the raw brain MRI images that were generated using an image-process pipeline and quality controlled centrally by the UKB (Alfaro-Almagro et al. 2018). In this work, we included subcortical (of the accumbens, amygdala, caudate, hippocampus, pallidum, putamen, and thalamus), total brain, and white matter hyperintensity volumes. All subcortical volumes were taken as the average between the left and right measures and adjusted for total intra-cranial volume. Total brain volume was adjusted for head size. All measures had units mm^3^. In the case of white matter hyperintensity volumes, whose distribution is left-skewed, we log-transformed these data. Finally, all brain volumes were clipped to enclose all values within 3-SDs of the mean. This yielded a maximum of 48,296 participants with genomic and segmented MRI data after individual and cohort quality control were performed on the data.

#### Neurological outcomes: all-cause, vascular, and Alzheimer’s dementia

Dementia was derived from the algorithmically determined outcomes (ADOs) produced by (Wilkinson et al. 2018) who used electronic health records to derive robust phenotypes for dementia (all-cause, vascular, and Alzheimer’s) in UKB participants. Briefly, these ADOs used curated codelists for various outcomes applied to hospital admissions, self-report, and death certificate data. We consider diagnoses from any sources. After applying individual and cohort quality control to the data, there were, respectively, 8,261 (2.2%), 1,789 (0.5%), and 3,726 (1.0%) cases of all-cause, vascular, and Alzheimer’s dementia out of 375,078 participants.

### Statistical methods

All analyses were performed using R version 4.3.1, PLINK 2.0 (Chang et al. 2015), the TwoSampleMR R package version 6.2.2 (Hemani, Tilling & Davey Smith 2017) (Hemani et al. 2018), and the Multivariable MR Package (Sanderson 2021). The multivariable MR method (MVMR) and package of (Sanderson 2021) was used to derive direct effects of amino acid levels and HbA1c (our chosen glycaemic mediator) on brain volumes and dementia status. This is illustrated in Figure 1 as a directed acyclic graph (DAG).

**Figure 1.**
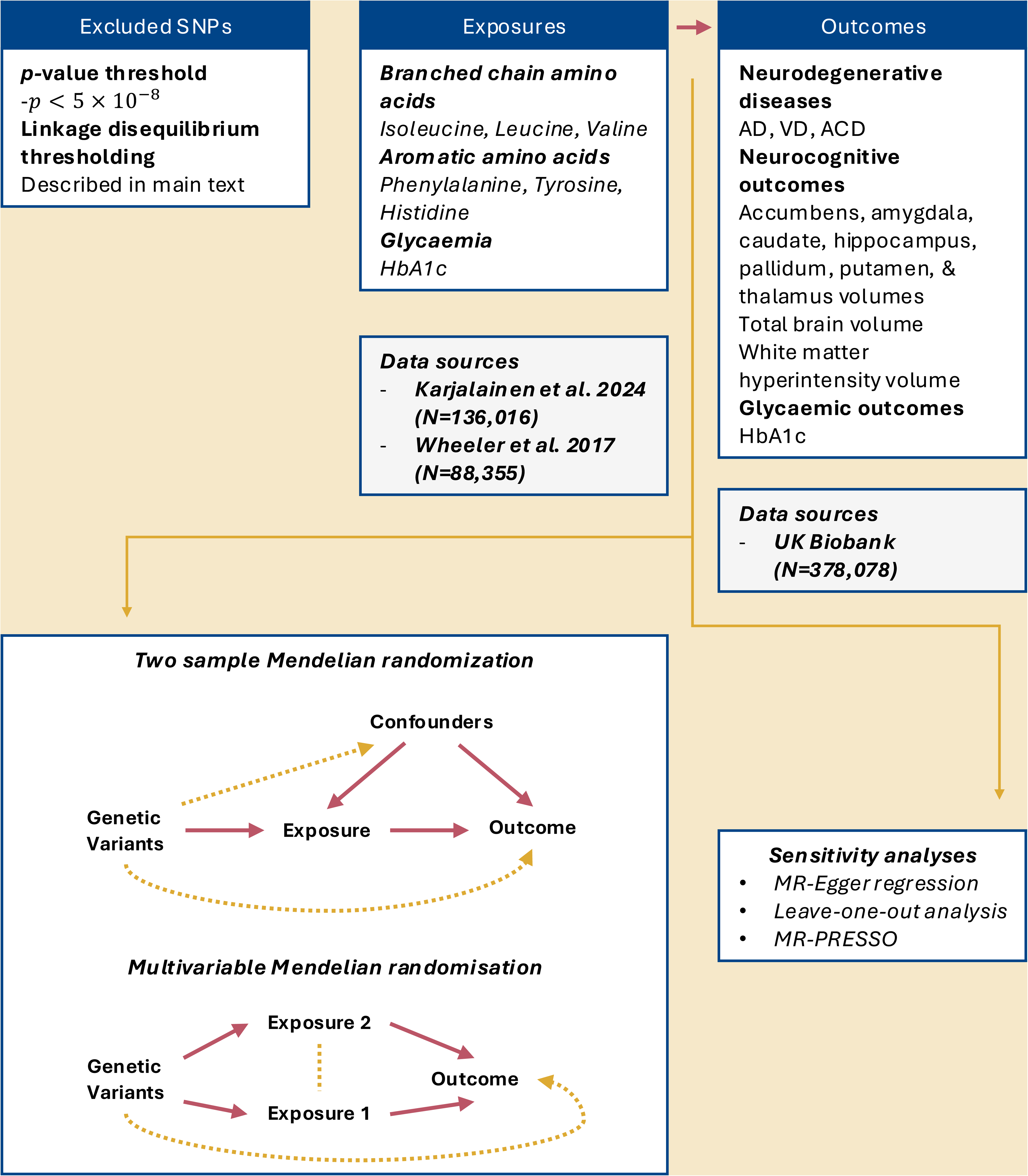
schematic illustrating study design.

#### Two sample MR analyses

We performed linear/logistic regressions to quantify the associations between the genetic instruments and outcomes in PLINK 2.0. All associations were adjusted for the first 10 genetic principal components to account for residual population stratification. Univariate estimates were obtained by combining the ratio of SNP–outcome to SNP–exposure associations across variants using a fixed-effects approach. Multivariable Mendelian randomization was then used to estimate the direct effects of HbA1c and circulating amino acids on each outcome. This approach jointly models the SNP–exposure associations for both traits so that each effect is estimated conditional on the other.

We also performed additional univariate MR sensitivity analyses, including MR-Egger regression (which allows for the *y*-intercept as a free parameter rather than zero, where the former would indicate unbalanced horizontal pleiotropy), the weighted median estimator (WME – which may yield robust estimates in cases where 50% of the genetic variants are invalid), and MR-Pleiotropy RESidual Sum and Outlier (MR-PRESSO; which can detect and correct for horizontal pleiotropy on-the-fly via removal of detected outlier SNPs) methods. Sensitivity analyses for the MVMR modelling included measurement of the instrument heterogeneity. Conditional instrument strength was also assessed by computing the F-statistic.

#### MR assumption checks

MR has three strict assumptions that must be met for study results to be valid:

I. *Relevance* – genetic variants being used as an instrument should be robust and strongly associated with the exposure of interest. Our exposure data were derived from a published large-scale GWAS (Karjalainen et al. 2024), thus satisfying this requirement.
II. *Independence* – there is no confounding of the association between the instrumental variable (IV) and the outcomes, for example by population stratification, assortative mating, and confounders of the SNP-outcome association. Population stratification was accounted for by including the first ten genetic principal components as covariates when computing SNP-outcome associations.
III. *Exclusion* – there is no independent pathway linking the instrument and the outcome of interest, other than via the exposure. We implemented three standard sensitivity analyses in our study: Egger regression, the weighted median (WME) estimator, and MR-PRESSO.

#### Presentation of results and transformations

Results are shown as multi-panel forest plots with an outcome per panel e. Figs. 2 and 3 show the MR estimates for continuous neuroimaging outcomes that were not log-transformed (they were normally distributed), whereas Figs. 4 and 5 show estimates for clinical outcomes (all-cause, vascular, and Alzheimer’s dementia) in addition to exponentiated point estimates for white matter hyperintensity volume on the same scale.

**Figure 2.**
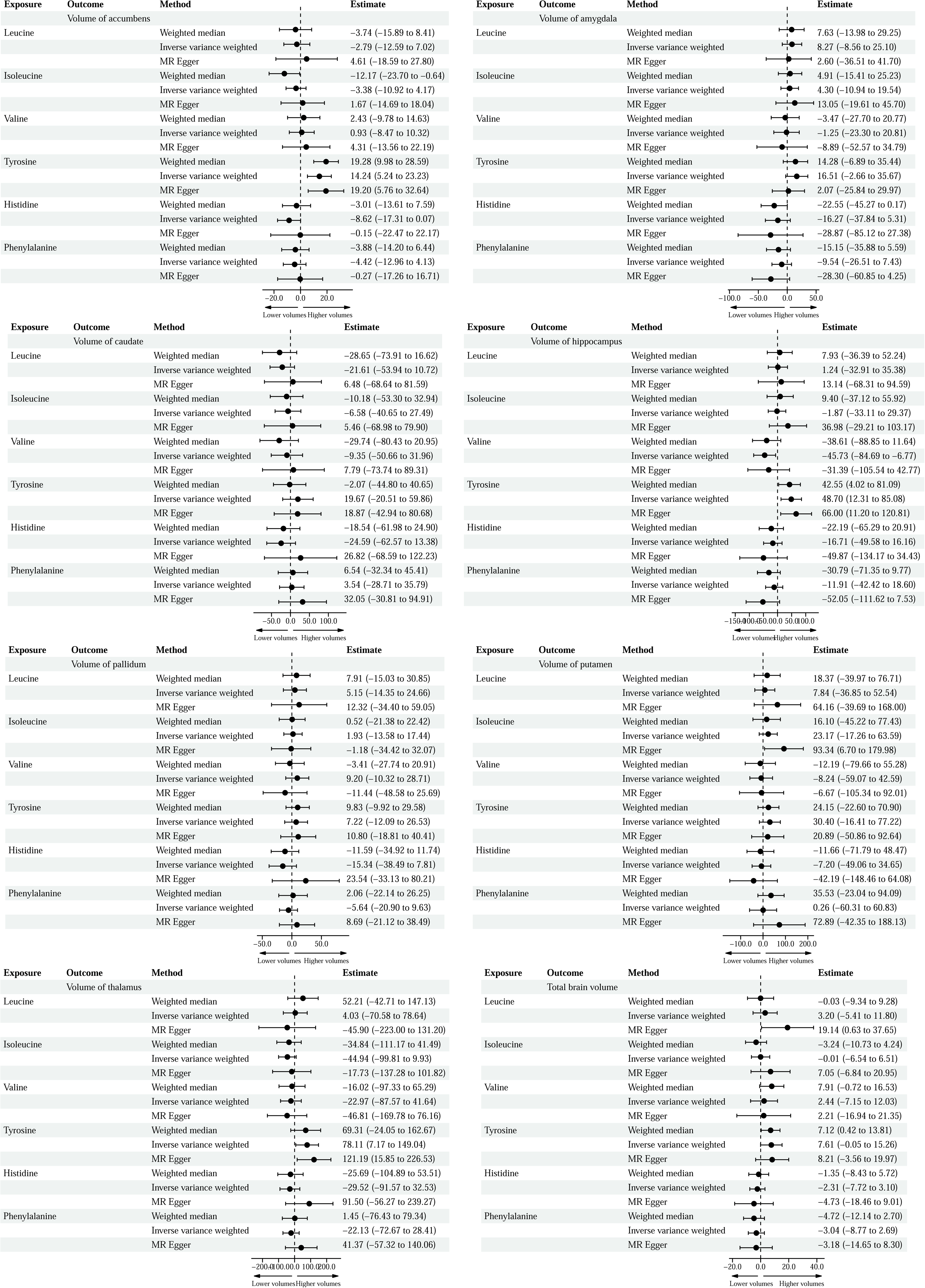
Univariate associations of BCAAs (leucine, isoleucine, valine) and AAAs (tyrosine, histidine, phenylalanine) with subcortical and total brain volumes in the UK Biobank.

**Figure 3.**
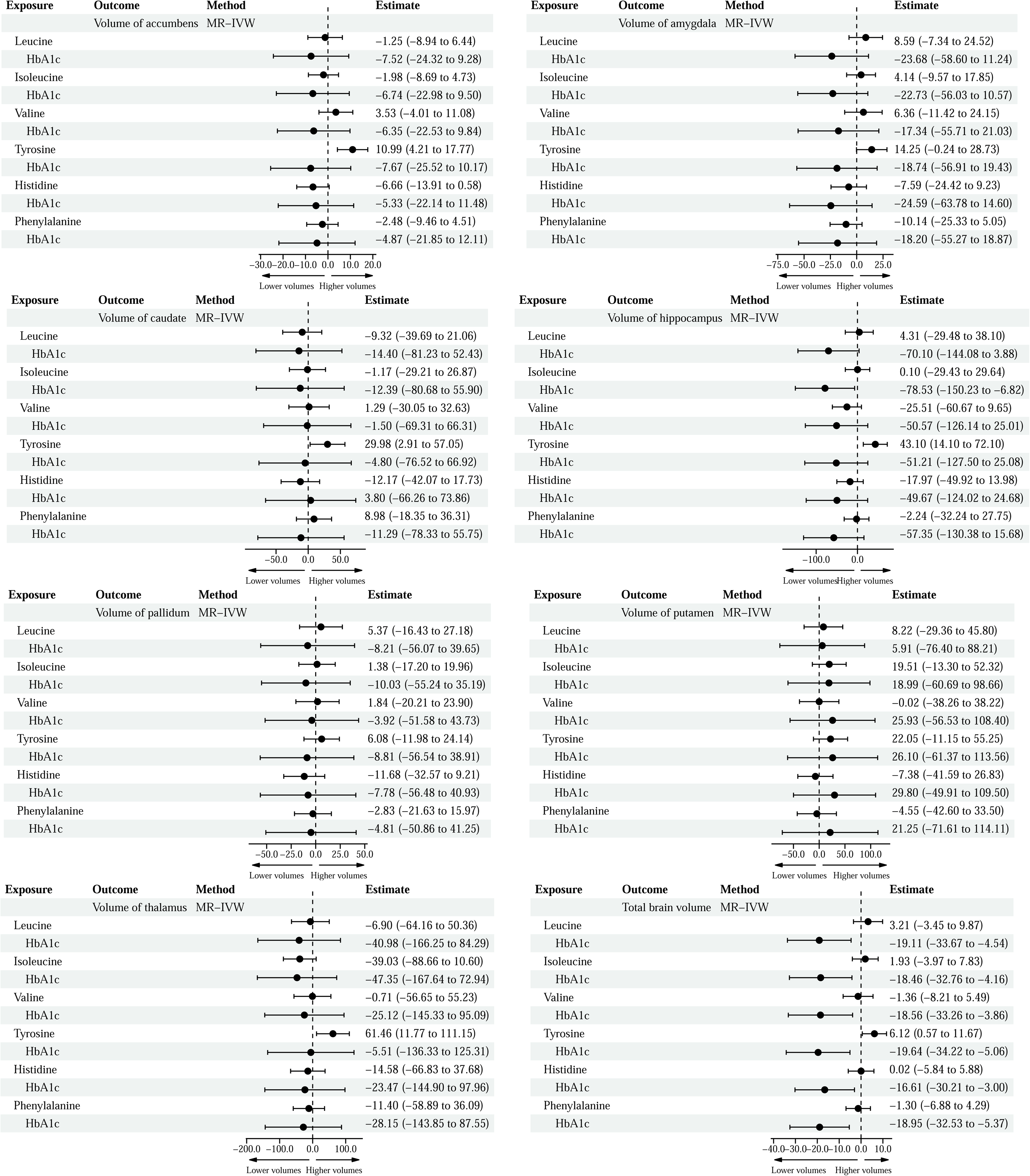
direct effects of BCAAs (leucine, isoleucine, valine), AAAs (tyrosine, histidine, phenylalanine), and glycaemia (HbA1c) on subcortical and total brain volumes in the UK Biobank from MVMR analyses.

**Figure 4.**
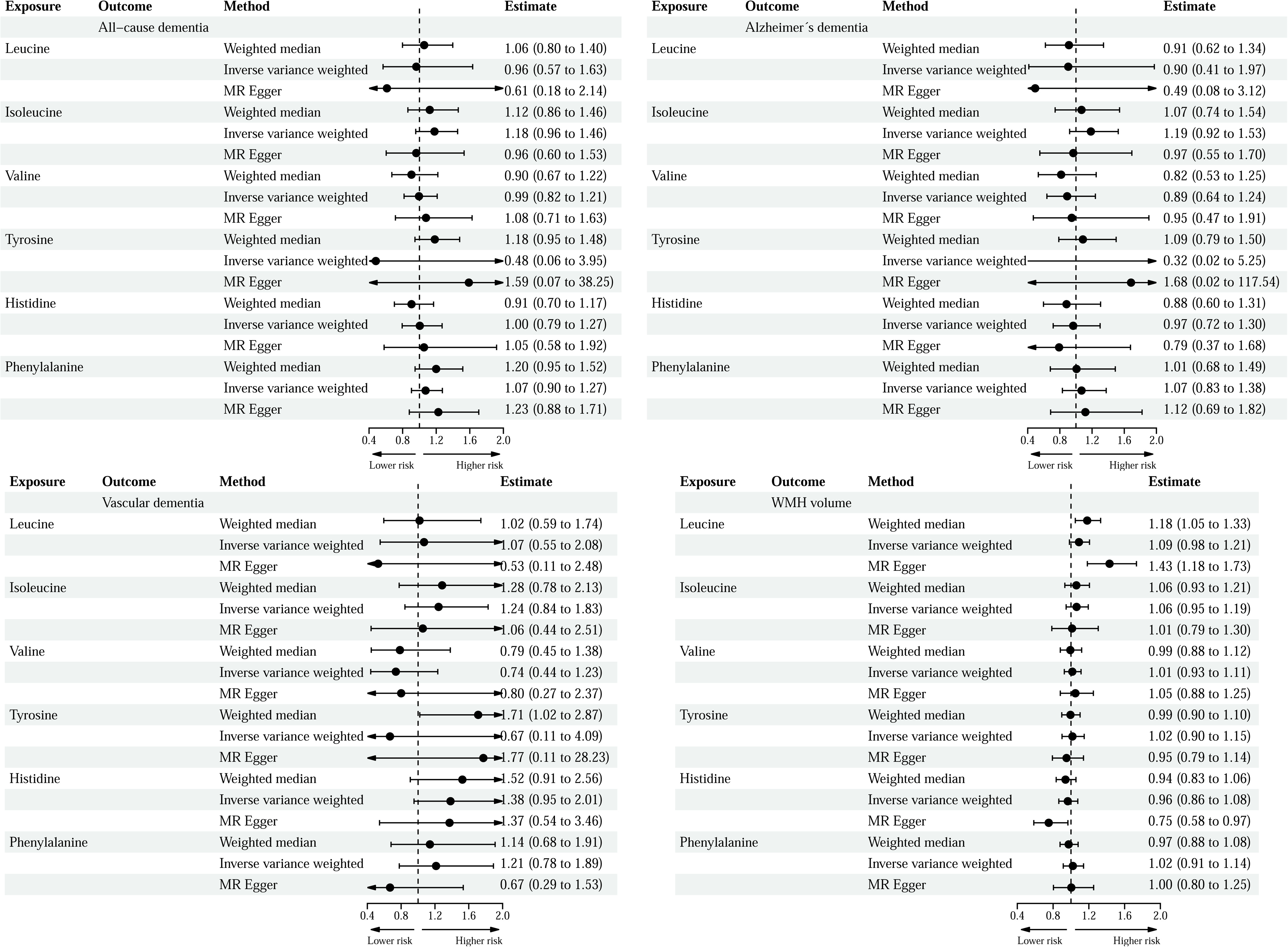
univariate MR associations for dementia and white matter hyperintensity volumes in the UK Biobank.

**Figure 5.**
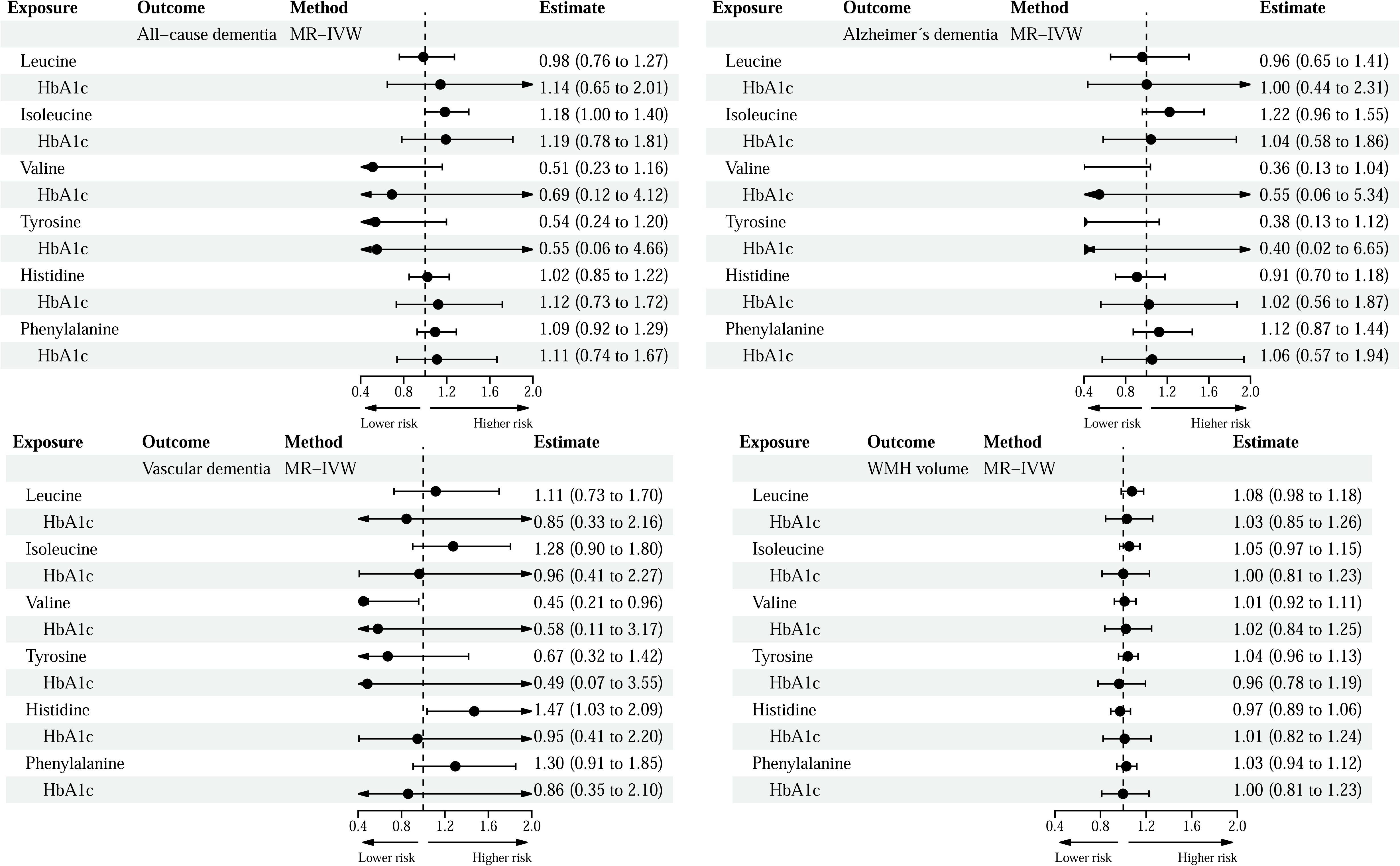
direct effects of BCAAs, AAAs, and glycaemia on dementia and white matter hyperintensity volumes in the UK Biobank.

## Results

The overall sample comprised up to 375,078 participants, with a mean age of 56.9 years. There were more females (54.1%). The mean circulating amino acid concentrations (in units mmol/L) were 0.05, 0.10, 0.21, 0.06, 0.07, and 0.05 for isoleucine, leucine, valine, tyrosine, histidine, and phenylalanine, respectively. The mean HbA1c level was 36.0 mmol/mol, and the mean BMI was 27.1 kg/m^2^. Characteristics of the sample are summarised in Table 1, including circulating amino acid levels.

### Associations between BCAA levels, HbA1c, and brain health

Univariate analyses of the branched chain amino acids (Fig. 2; leucine, isoleucine, and valine) showed a negative association between genetically proxied valine and hippocampal volume (β = -45.7 95%CI [-84.69, -6.77] mm^3^/mmol L^-1^). The MVMR direct estimate (Fig. 3) crossed the null (β = -25.51 95%CI [-60.67, 9.65] mm^3^/mmol L^-1^) but was directionally consistent suggesting that this association may be partially mediated by glycaemia. No other associations between branched chain amino acids and subcortical, total brain, or white matter hyperintensity volumes were observed.

We found evidence for a risk-lowering effect of elevated valine levels on vascular dementia (Fig. 5; OR = 0.45 95% CI [0.21, 0.96] / mmol L^-1^) in our MVMR analysis that was independent of HbA1c. By contrast, the univariate IVW point estimate (Fig. 4) was OR = 0.74 95% CI [0.44, 1.23] / mmol L^-1^. No other associations were found.

### Associations between AAA levels, HbA1c, and brain health

Univariate MR analyses of the aromatic amino acids (Fig. 2; phenylalanine, tyrosine, and histidine) revealed positive associations between elevated circulating tyrosine and the volumes of the accumbens (β = 14.24 95%CI [5.24, 23.23]), hippocampus (β = 48.7 95% CI [12.31, 85.1]), and thalamus (β = 78.1 95% CI [7.17, 149.0]). MR PRESSO identified SNP rs12206654 as an outlier in the estimate between tyrosine and the volume of the thalamus, resulting in an outlier-corrected estimate of β = 15.8 95% CI [-56.7, 88.2]. This suggests that this association is driven by pleiotropy.

MVMR estimates (Fig. 3) revealed a direct effect of tyrosine on the volumes of the caudate (β = 30.0 95% CI [2.91, 57.05]) and total brain volume (β = 6.12 95% CI [0.57, 11.67]). The latter was independent of the total and direct effect of HbA1c on total brain volume seen in univariate and MVMR estimates (β = -18.3 95%CI [-33.4, -4.5]).

Univariate MR estimates for circulating AAAs on dementia and white matter hyperintensity volumes yielded no associations (Fig. 4). However, we find evidence of an association between histidine and increased vascular dementia risk (OR = 1.47 95% CI [1.03, 2.09]) that was independent of HbA1c. No other associations were observed.

## Discussion

### Summary of main findings

In this large-scale population-based study, we investigated whether there is a causal relationship between elevated amino acid levels and brain health independent of glycaemia for a maximum of *N*=375,078 (54.9% female) UK Biobank participants.

Using multivariable MR with an instrument for HbA1c, we showed that elevated tyrosine levels may be associated with enhancements of some subcortical volumes. These effects were independent of HbA1c suggesting no mediation by glycaemia. Valine and histidine were also associated with lower and higher risks of vascular dementia, respectively. Valine was also associated with lower hippocampal volume, although this result was attenuated in the MVMR estimate, demonstrating that this may be mediated by glycaemia.

### Interpretation in the context of other research

Our results suggest that elevated tyrosine levels could have beneficial effects on brain health. This is in agreement with multiple studies and trials that have demonstrated that acute administration or supplementation of tyrosine results in improvements in cognitive performance (Deijen et al. 1999; Roiser et al. 2005; Van De Rest et al. 2017). However, animal studies historically have shown that chronically elevated tyrosine is associated with the development of metabolic dysregulation, sensory neuropathies, and DNA damage to the brain (Boctor & Harper 1968; De Prá et al. 2014; Dickinson et al. 2004; Nagaoka et al. 1986). Such outcomes also feature in tyrosinemia (a rare inborn error of tyrosine metabolism) in humans although it is important to note that levels of tyrosine characteristic of tyrosinemia can be at least four-fold higher than in healthy controls (Sikonja et al. 2022). Furthermore, there is evidence of an age and dose-dependent effect of tyrosine intake, with older adults experiencing domain-specific adverse cognitive effects (Bloemendaal et al. 2018; Kühn et al. 2019).

Parkinsonism is a rare complication of PKU – it has been speculated that chronically elevated phenylalanine results in a depletion of tyrosine in the brain, inhibiting dopamine production and results in PD symptoms such as tremor and mood swings (Manti et al. 2026; Tufekcioglu et al. 2016; Velema et al. 2015). Imaging studies have demonstrated a correlation between incident PD and reduced volumes of the caudate, amygdala, putamen, and thalamus (Colpan & Slavin 2010; Lee et al. 2011; Lisanby et al. 1993; O’Neill et al. 2002). Elevated circulating levels of O-sulfo-L-tyrosine, a derivative of tyrosine, have also been shown to be correlated with a reduced risk of incident PD in a previous two-sample MR study (Liao et al. 2025).

Elevated valine has been associated with a decreased dementia risk observationally using UKB data (Fu et al. 2024). Bidirectional MR analyses have suggested that reduced valine concentrations are secondary to Alzheimer’s disease, and it has been suggested as a biomarker for early diagnosis (Qian et al. 2023). Valine levels have also been demonstrated to decline as a function of age in vascular dementia (Ma et al. 2025), which may also be the case in Alzheimer’s dementia according to the results presented here.

Elevated histidine has been associated observationally with a reduced risk of VaD (Qiang et al. 2024), in contrast to the association reported here between elevated histidine and higher risk of VaD. However, this apparent discordance may reflect limitations inherent to observational study designs. The protective association observed by Quiang et al. may reflect reverse causation in the observational setting whereby individuals with VaD may present with reduced circulating histidine. This would be consistent with the broad association between depleted aromatic amino acids and VaD (Aquilani et al. 2023). However, our finding that elevated histidine may increase VaD risk may be biologically plausible. Rodent studies investigating the effects of low protein diets on metabolic health have demonstrated that restricting dietary histidine can improve body composition and glycaemic biomarkers (Flores et al. 2023). Taken together with the histidine→VaD MR estimate, it is possible that elevated circulating histidine may adversely affect metabolic pathways implicated in the pathophysiology of VaD, warranting further investigation. It is worth commenting on the apparent discordance between the valine→hippocampus association measured in UKB, which was negative. Progressive hippocampal atrophy has been well-documented in dementia (Raket et al. 2026), yet for valine we measured directionally discordant associations and hippocampal volume (negative) and VaD (risk-decreasing). The attenuated estimate for this association in Fig. 3 from our MVMR analyses demonstrated that the former association may be mediated by glycaemia. In addition, although hippocampal atrophy has been demonstrated in VaD this depends on the stage and severity of the disease. Prior imaging studies have demonstrated similar heterogeneity in estimates of the association between subcortical volumes and dementia risk (Rosbergen et al. 2025; Van Der Velpen et al. 2023).

### Strengths

UKB is the largest imaging study undertaken, with segmented brain scans available for almost 100,000 participants. Clinical outcomes like dementia and T2DM were algorithmically defined leveraging both self-reported and electronic healthcare record data to produce robust outcomes.

The Mendelian randomisation approach evaluates sustained exposure to these elevated amino acid levels over the life-course. These associations could offer additional insight into the underlying pathogenesis of neurological disorders and brain atrophy, as well as possible diet and supplementation strategies to minimise the risk of dementia in later life.

### Limitations

There are some limitations worthy of discussion. An important caveat to these results is in the sample selection. We limited our analyses to individuals of white British ancestry, and these associations may differ in individuals of different ancestries. Performing the same analysis *ab initio* for other genetic ancestries may reveal markedly different associations.

While chronically elevated circulating amino acid levels are correlated with CSF amino acid levels, the latter would be a better proxy of amino acid levels in brain tissue and perhaps yield more robust estimates. However, CSF metabolites would require that UKB participants undergo lumbar puncture (LP) to extract samples for metabolites to be measured, a far more invasive procedure to obtaining plasma samples for the NMR metabolomics data in UKB.

Though we performed several sensitivity analyses to complement our main analysis, Mendelian randomisation fundamentally assumes that the genetic instruments adopted are associated with the outcome strictly via the risk factor of interest. This was mitigated somewhat using MVMR, MR-PRESSO, and MR-Egger analysis which indicated that the estimates were largely not affected by widespread pleiotropy except for the association between tyrosine and thalamus volume. Another caveat is that when framed in terms of the proportion of the variance in AA levels and HbA1c explained by each instrument, only a small fraction of the variation in amino acid and HbA1c levels are explained by the instruments.

A two-step MR framework could, in principle, quantify the proportion of the AA-brain association mediated by glycaemia (i.e., HbA1c). However, the absence of strong independent BCAA→HbA1c and HbA1c→brain associations renders the product of coefficients approach typically adopted in the literature uninformative due to compounding uncertainty. Hence, MVMR was adopted to address whether any associations between circulating AAs and brain volume were independent of glycaemia.

### Unanswered questions and future directions

The present analyses cannot inform the causal ordering between circulating AA concentrations and glycaemia. Bidirectional MR, where instrumental variables for the outcomes presented in this paper would be used as exposures and metabolites the outcomes, is required to determine whether the attenuation of some AA→brain associations reflect that elevated AA concentrations are secondary to *i)* glycaemia, or *ii)* brain atrophy and dementia.

Not all the large essential AAs have been assayed in the UKB data. Tryptophan, the precursor to melatonin and serotonin, has been shown to be positively associated with sleep, mood, cognition, and brain health (Jenkins et al. 2016; Silber & Schmitt 2010) in addition to being implicated in the pathogenesis of Alzheimer’s disease (Savonije & Weaver 2023). It is therefore plausible to expect a positive association between neuroimaging and neurological outcomes and tryptophan (Henry et al. 2019; Spira et al. 2014). However, to date, few loci associated with tryptophan have been identified in GWASes, and a formal assessment of instrument strength would be difficult without tryptophan measurements included in the data. This is a significant limitation as we are unable to robustly account for the large essential aromatic amino acids.

The instrument comprised SNPs associated with circulating random amino acid levels, meaning that the associations reflect SNP-exposure effects calculated using likely a mixture of postabsorptive and postprandial data. The same applies to the circulating plasma AA levels recorded in UKB. Whether postprandial AA excursions, which may engage different distinct pathways, exert independent effects on brain structure or dementia risk cannot be addressed within the current framework, and represents an important direction for future investigations if large-scale postprandial metabolomic GWASes become available.

### Implications for clinical and regulatory science

Recent MR studies also revealed that excess circulating tyrosine is associated with shorter lifespan in males (Zhao et al. 2025). Thus, balancing the benefits and risks of acute tyrosine administration must be done in the context of results replicating the main MR analyses here; as well as observational, and future experimental studies quantifying the longitudinal effect of excess dietary tyrosine in humans. Since the metabolites considered in this study are frequently measured in population-scale cohorts similar to UKB, we aim to reproduce these results in different cohorts where there is no overlap with the GWAS study of (Karjalainen et al. 2024), such as in FinnGen (Kurki et al. 2023).

In conclusion, we demonstrated for the first time using MR that elevated tyrosine is associated with increases in some subcortical volumes that have been previously associated with the onset and progression of multiple neurological disorders. For valine and histidine, we found evidence of causal associations with reduced and increased risk of vascular dementia, respectively, which must be investigated in future work. Amino acid levels are a modifiable exposure that can easily be intervened on via redistribution, restriction or enhancement of protein intake, and acute administration.

## Data Availability

This research has been conducted using the UK Biobank Resource under Application Number 422316. This work uses data provided by patients and collected by the NHS as part of critical care and support. Data can be accessed by approved researchers through the UK Biobank Access Management System.

## Data availability

Data are available through the UK Biobank (http://www.ukbiobank.ac.uk/) upon application and with permission of UKBB’s Research Ethics Committee.

## Acknowledgements

This research has been conducted using the UK Biobank Resource under Application Number 422316. This work uses data provided by patients and collected by the NHS as part of critical care and support. We gratefully acknowledge the UK Biobank participants for their contributions, without whom this research would not have been possible.

## Supplementary material

Supplementary material (Tables S1-S) available online.

## Abbreviations

No abbreviations list. Abbreviations should be defined at first use (except for accepted gene/protein symbols). Abbreviations in figures and tables should be defined in the legend.

The following do not need defining: AIDS; ANOVA; ATP; cDNA; CNS; CSF; CT; DNA; ECG; EEG; ELISA; EMG; GABA; HIV; MRI; PBS; PCR; PET; REM; RNA; mRNA; tRNA

